# Head and body dyskinesia during gait in tactical athletes with vestibular deficit following concussion

**DOI:** 10.1101/2021.04.27.21256200

**Authors:** John J. Fraser, Jacob VanDehy, Dawn M. Bodell, Kim R. Gottshall, Pinata H. Sessoms

**Author notes:** Corresponding author: John J Fraser, Warfighter Performance Department, Naval Health Research Center, 140 Sylvester Road, San Diego, CA 92106, USA., Twitter: @NavyPT. Disclaimer: The author(s) are military service members or employees of the U.S. Government and this work was prepared as part of their official duties. Title 17, U.S.C. §105 provides that copyright protection under this title is not available for any work of the U.S. Government. Title 17, U.S.C. §101 defines a U.S. Government work as work prepared by a military service member or employee of the U.S. Government as part of that person’s official duties. Report No. 19-69 was supported by the U.S. Navy Bureau of Medicine and Surgery under work unit no. N1703. The views expressed in this article are those of the authors and do not necessarily reflect the official policy or position of the Department of the Navy, Department of Defense, nor the U.S. Government. The study protocol was approved by the Naval Health Research Center Institutional Review Board in compliance with all applicable Federal regulations governing the protection of human subjects. Research data were derived from an approved Naval Health Research Center Institutional Review Board protocol, number NHRC.2015.0010. All participants provided informed consent. Presented at the Military Health System Research Symposium, August 20, 2018, Orlando, FL and is archived at doi:10.7490/f1000research.1115992. The preprint of the manuscript is archived on medRxiv and accessible at doi:10.1101/2021.04.27.21256200.

## Abstract

**Background:** Vestibular deficit is common following concussion and may affect gait. The purpose of this study was to investigate differences in head and pelvic center of mass (COM) movement during gait in tactical athletes with and without concussion-related central vestibular impairment.

**Material & Methods:** 24 patients with post-concussion vestibular impairment (20 males, 4 females; age: 31.7±7.9 years; BMI: 27.3±3.3) and 24 matched controls (20 males, 4 females; age: 31.8±6.4 years; BMI: 27.2±2.6) were included in the analyses. Three-dimensional head and pelvic COM displacement and velocities were collected at a 1.0 m/s standardized treadmill walking speed and assessed using Statistical Parametric Mapping t-tests. Maximum differences (d_max_) between groups were reported for all significant kinematic findings.

**Results:** The Vestibular group demonstrated significantly diminished anteroposterior head excursions (d_max_=2.3 cm, p=0.02;) and slower anteroposterior (d_max_=0.37 m/s, p=0.01), mediolateral (d_max_=0.47 m/s, p=0.02) and vertical (d_max_=0.26 m/s, p<0.001) velocities during terminal stance into pre-swing phases compared to the Control group. Vertical pelvic COM excursion was significantly increased in midstance (d_max_=2.4 cm, p=0.03) and mediolaterally during pre-to initial-swing phases (d_max_=7.5 cm, p<0.001) in the Vestibular group. In addition, Pelvic COM velocities of the Vestibular group were higher mediolaterally during midstance (d_max_=0.19 m/s, p=0.02) and vertically during post-initial contact (d_max_=0.14 m/s, p<0.001) and pre-swing (d_max_=0.16 m/s, p<0.001) compared to the Control group.

**Significance:** The Vestibular group demonstrated a more constrained head movement strategy during gait compared with Controls, a finding that is likely attributed to a neurological impairment of visual-vestibular-somatosensory integration.

## 1. Introduction

Concussion, also known as mild traumatic brain injury (mTBI), is a common clinical entity experienced by more than 22 400 tactical athletes in the United States Armed Forces(1) and is responsible for more than 2.8 million civilian emergency room visits per annum.(2) Many individuals who sustain a concussion experience signs and symptoms that include dizziness, imbalance, dyskinesis, and cognitive deficit.(3) These manifestations are a result of neurophysiological impairment in higher-order brain function, pyramidal and extrapyramidal motor pathways, and vestibular function.(4) These alterations in visual-vestibular-somatosensory integration may persist well beyond the initial injury. Athletes with prior history of concussion have been found to have lasting alteration in static and dynamic postural control at an average 44 months following injury.(5) Visual-vestibular-somatosensory integration deficits may affect smooth pursuit, vestibulo-ocular reflex (VOR), dynamic visual acuity, gaze stabilization, subjective visual vertical, and spatial orientation during execution of complex motor tasks such as walking.

Individuals with concussion have been reported to have gait differences compared with healthy controls. In a recent systematic review assessing gait changes following concussion, individuals have been found to walk slower in the early acute phase (6 of 14 studies) that resolved within 10 days (10 of 13 studies).(6) There were mixed results for measures of stride length acutely (decreased in 2 of 7 studies) and double support time (increased in 2 of 5 studies) from intermediate to long-term time-points following injury. (6) Decreased anteroposterior (4 of 13 studies) and increased mediolateral center of mass (COM) movement (3 of 12 studies) was observed acutely following injury that was primarily resolved by 10 days post-injury (8 of 9 studies).(6) It is highly likely that the wide variability of findings observed during gait is related to heterogeneity of neurological impairment found in this clinical population.

Little is known regarding the effects of central vestibular impairment on head and pelvic kinematics during walking following concussion. In a preliminary study of head and trunk mechanics during gait, tactical-athletes with vestibular deficit resulting from concussion were found to have asynchrony and large variability of the overall head position in relation to the pelvic COM.(7) These findings warrant further investigation due to the small sample size and large observed variability.(7) Therefore, the purpose of this study was to investigate head and trunk mechanics during gait in tactical-athletes with and without vestibular deficit following concussion.

## 2. Material & Methods

A descriptive laboratory cross-sectional study was performed where the independent variable was group (Control, Vestibular). The outcome measures were COM displacement and velocity of the head and pelvis in the sagittal, frontal, and transverse planes during treadmill walking at a standardized 1.0 m/s.

### 2.1. Participants

After excluding one participant for suspected secondary gain, 24 tactical athletes in the United States Armed Forces with post-concussion vestibular impairment (20 males, 4 females; age: 31.7±7.9 years; BMI: 27.3±3.3) recruited from a military vestibular rehabilitation clinic were included in the analyses. The control group was comprised of 24 tactical athletes without a history of concussion (20 males, 4 females; age: 31.8±6.4 years; BMI: 27.2±2.6) that were matched on sex, age, and BMI (Table 1). Participants in both groups were included if they were active duty military between the ages of 18 and 50. Those who incurred a concussion 6 to 52 weeks prior to consent and had a medical diagnosis of a concussion-related central vestibular disorder were included in the post-concussion Vestibular group [Dizziness Handicap Inventory: 41.0±19.9; Activities-specific Balance Confidence Scale: 73.7±17.1; Functional Gait Assessment: 23.8±4.5; Computerized Dynamic Posturography Sensory Organization Test (SOT) Composite: 63.1±12.1]. Healthy controls must have passed their most recent physical fitness assessment.

**Table 1.** Group demographics and spatiotemporal gait characteristics.

|  | Control<br>(n=24)<br>20 males, 4 females | Vestibular<br>(n=24)<br>20 males, 4 females |  |
| --- | --- | --- | --- |
| | Mean ± SD | | *p*-value |
| Age (years) | 31.8 ± 6.4 | 31.7 ± 7.9 | .68 |
| Height (cm) | 175.2 ± 9.8 | 174.8 ± 9.1 | .76 |
| Weight (kg) | 83.8 ± 12.4 | 83.9 ± 13.8 | .96 |
| BMI (kg/m<sup>2</sup>) | 27.2 ± 2.6 | 27.3 ± 3.3 | .84 |

Participants were excluded from participation if they had an orthopaedic condition that could affect gait, impaired joint mobility, benign positional vertigo, fluctuating peripheral vestibular dysfunction, non-organic behavior, conversion reactions, peripheral neuropathy, oculomotor nuclei (III, IV, VI) abnormalities, motor impairment, central neurological diseases, cerebral vascular accident, or were pregnant. Each participant gave informed consent before participating in the study. While participants did not directly contribute to the design of this study, specialist clinicians who care for and advocate for this patient population contributed. Regarding dissemination, study participants will be provided a link to a website that explains the research findings in common language. This study was approved by the Naval Health Research Center Institutional Review Board (protocol NHRC.2015.0010).

### 2.2. Procedures

Figure 1 details the study flow sheet from recruitment to analysis. Following consent, patients provided demographic information, and health and injury history. Height and mass were measured. Patients were asked to walk at a 1.0 m/s pace on a dual-belt instrumented treadmill (Motekforce Link, Amsterdam, The Netherlands) while looking forward at an optotype placed on a screen in front of the treadmill. Subjects wore a safety harness to prevent them from falling but the harness did not provide weight support.

**Fig. 1.**
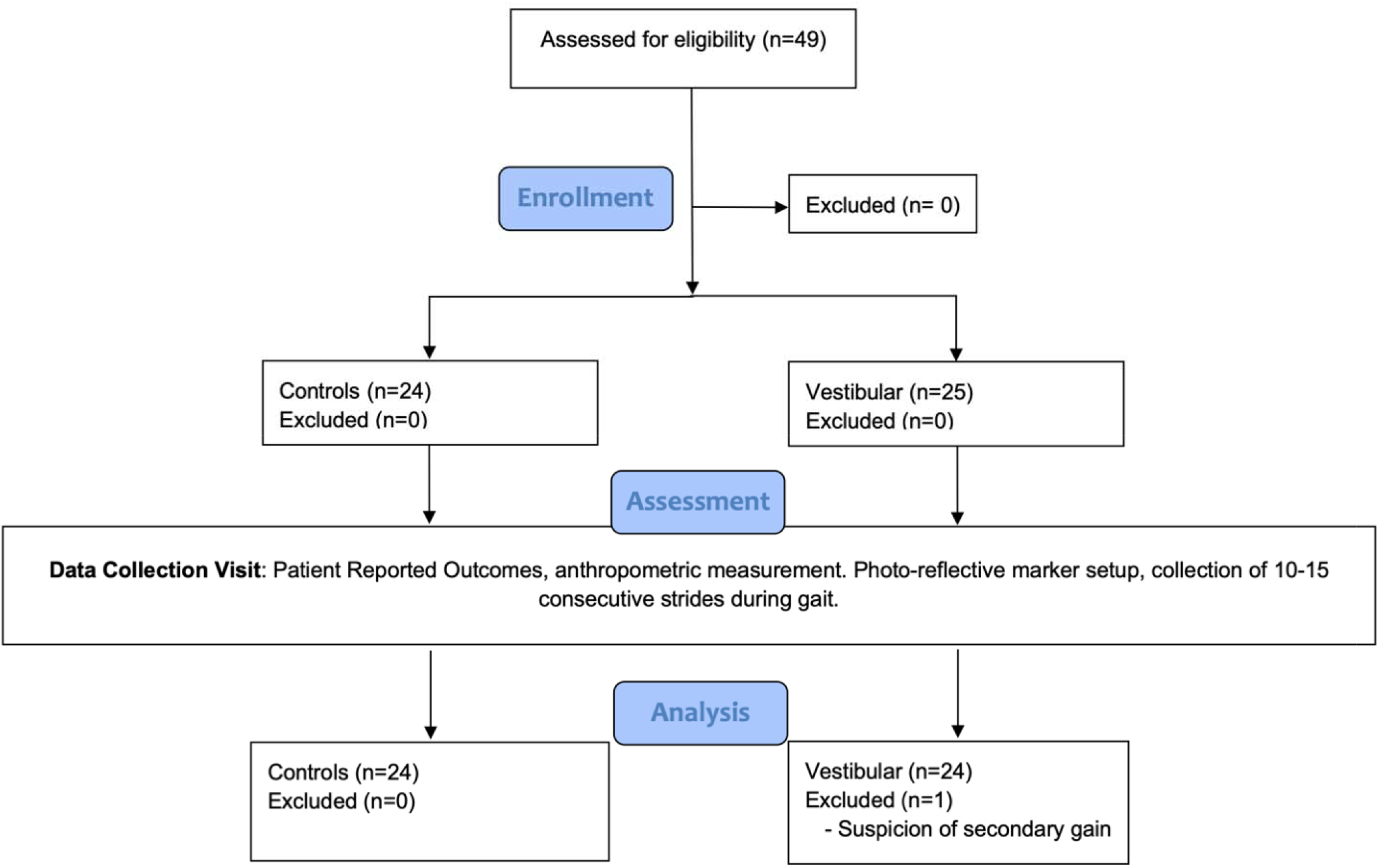
Study flow diagram.

Kinematic data were collected with a 14-camera optical motion capture system (Motion Analysis Corp., Santa Rosa, CA, USA) at 120 Hz and referenced to the world. Three-dimensional measurements of COM displacement and velocity for 10–15 consecutive strides were averaged using 101 data points and normalized to initial contact (IC). Head movement was tracked using a total of five photo-reflective markers affixed to a fitted ball cap worn on the head (left, front, right, back, and top of the head). A cluster of four markers affixed to a rigid plastic fixture and placed on the sacrum measured pelvic motion. A single heel marker affixed to the counter of each shoe were used to demarcate initial contact during gait. Kinematic data were processed using the Visual3D software (C-Motion Inc., Germantown, MD, USA). Head and pelvic COM were calculated as the average X,Y, Z position of the head and pelvic markers, respectively, while foot contact positions were collected using the heel markers. Velocities were calculated as the first derivative of the positional data.

### 2.3. Statistical analysis

An *a priori* sample size estimation of 16 participants were needed based on the variance of head stabilization,[6] an *α*=.05, and *β*=.20. Descriptive statistics were calculated for demographic and spatiotemporal gait characteristics and assessed for differences with independent t-tests using MATLAB (MathWorks, Inc., Natick, MA, USA). Group differences in triplanar motion of the head and pelvic COM over the gait cycle were assessed using Statistical Parametric Mapping (SPM) t-tests using spm1d version 0.4, a package written by Pataky(8) for one-dimensional SPM analysis for Python 3.6.5 (Python Software Foundation, Beaverton, OR, USA). Maximum differences (d_max_) and mean difference (d_mean_) between the two groups were reported for all significant kinematic findings.

## 3. Results

There were no significant differences between the Vestibular and Control groups for age, height, mass, or BMI (Table 1). The Vestibular group walked with a significantly wider base of support (Table 1). There were no other significant gait differences between groups. Figure 2 illustrates the COM motion of the head and pelvis in three dimensions. Several significant group differences were observed in head and pelvic COM movement during gait (Figure 3). The Vestibular group demonstrated diminished anteroposterior head excursions (d_max_= 2.3 cm, p = 0.02) and slower anteroposterior (d_max_= 0.34 m/s, p < 0.001), mediolateral (d_max_=0.47 m/s, p = 0.02) and vertical (d_max_= 0.26 m/s, p < 0.001) velocities during terminal stance into pre-swing phases compared to controls. Vertical pelvic COM excursion was significantly increased in midstance (d_max_= 2.4 cm, p = 0.03) and mediolaterally during pre-to initial-swing phases (d_max_= 7.5 cm, p < 0.001) in the Vestibular group. In addition, pelvic COM velocities were higher mediolaterally during midstance (d_max_= 0.19 m/s, p = 0.02) and vertically during post-IC (d_max_= 0.14 m/s, p<0.001) and pre-swing (d_max_= 0.16 m/s, p<0.001) in the Vestibular group compared to the Control group.

**Fig. 2.**
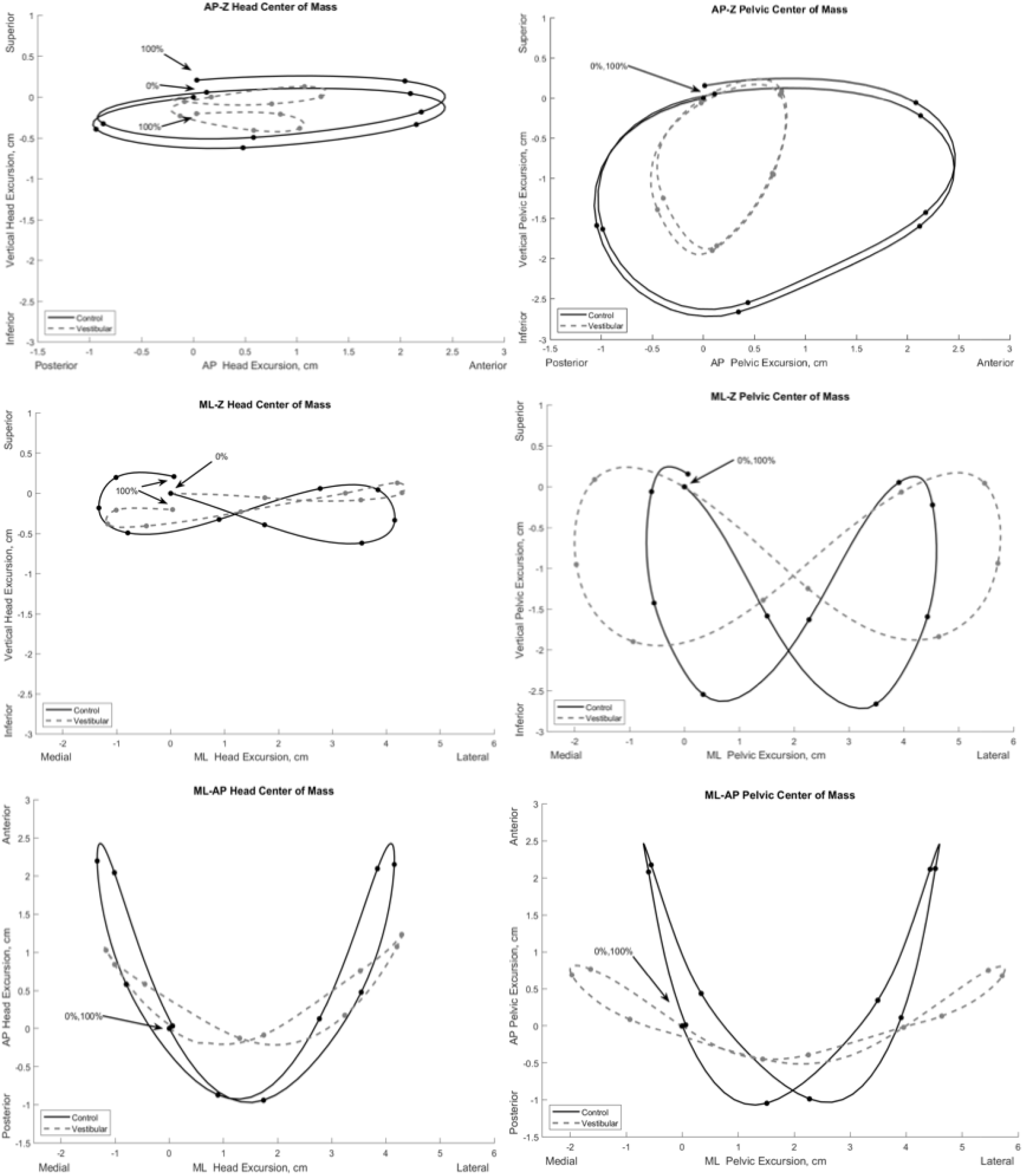
Mean three-dimensional excursion of the head and pelvic center of mass during gait in the Vestibular (dashed line) and Control (solid line) groups. Each point on the line depicts 10% increments of the gait cycle starting at initial contact (0%) to ipsilateral initial contact (100%). AP, anteroposterior; ML, mediolateral; Z, vertical.

**Fig. 3.**
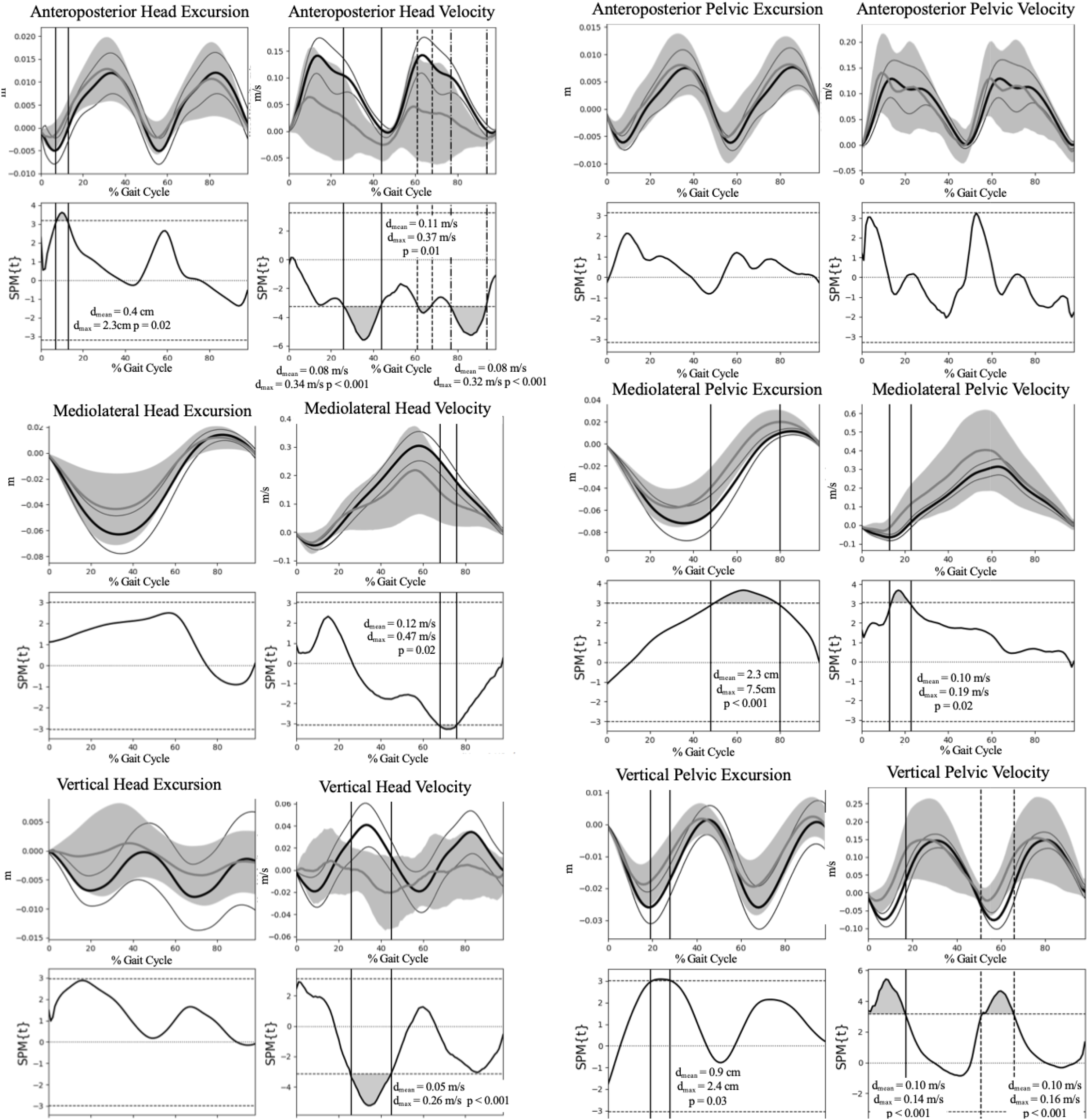
Group comparison of the head and pelvic center of mass excursion and velocity during gait. Group means and 95% confidence interval cloud for the Vestibular (grey mean line and confidence interval cloud) and Control (black mean line and white confidence interval cloud) groups are depicted. d_max_, maximum difference between groups; d_mean_, mean difference between groups; SPM, Statistical Parametric Mapping.

## 4. Discussion

The primary findings of this study were that patients with post-concussion vestibular deficit demonstrated a more constrained head but increased mediolateral and vertical pelvic movement strategy during gait compared with the Control group, a finding that is likely attributed to a deficit of the VOR, vestibulospinal reflex, and dynamic balance integration. To our knowledge, these findings are the first to be reported within this clinical subpopulation.

### 4.1. Head mechanics

We posit that the observed lower head excursion and velocity are likely a compensation for deficits in dynamic visual acuity (DVA), gaze stabilization, and subjective visual vertical affecting static and dynamic spatial orientation. From a kinematic perspective, movement of the head functions similarly to the proximal joints of the extremities. The mobility of the head allows for multiple degrees of freedom when exploring the environment visually. Similar to the extremities, proximal segmental stabilization of the head contributes to visual acuity, gaze stability, and spatial orientation. Vestibulopathy following concussion likely manifests with compensatory reduction of head excursion to try to stabilize gaze during dynamic gait. Further research incorporating optical tracking in relation to head movement during gait tasking is needed to substantiate this supposition.

The VOR, gaze stabilization, accurate subjective visual vertical, and the angular vestibulocollic (VCR) and cervicocollic (CCR) reflexes are important in head stabilization and control during static and dynamic function.(9) Increasing temporal stimulation of the semicircular canals, otoliths sensitive to gravity, and the eyes contribute to ‘velocity storage’ in the vestibular nuclei.(10) Individuals post-concussion may display high-gain VOR and a propensity for motion intolerance. Decreased excursion of head motion response to external moments is a countermeasure for the conflicting sensory afference when gait tasking is superimposed. Both the VCR and the CCR function to dampen head movement(9) and are normally inhibited during gait.(11) Our findings suggest that central vestibular impairment resulting from concussion may disinhibit these reflexes during walking,(9,12) resulting in diminished head movement during gait.

Mechanical or neurophysiological impairment resulting from zygapophyseal or muscular injury in the cervical spine may contribute to constrained head movement. Kinematics of concussion involved high accelerations of the head that are mitigated by cervical co-contraction.(13) Cervical spine pain is a common comorbidity following concussion, a condition that has many overlapping signs and symptoms.(14) It is plausible that injury to the cervical joints and muscles that serve an important role in providing afferent information regarding head position is disrupted. Furthermore, injury may change the contraction dynamics of the cervical musculature(9)and further increase the burden on impaired central control mechanisms.

Psychogenic factors are both plausible and a likely contributor to diminished head movement excursion and velocity following concussion. Individuals with psychological trauma resulting from the injurious event or other prior traumatic exposure may demonstrate altered movement strategies. Kinesiophobia, pain, and perceived function have previously been shown to be associated with altered cervical kinematics.(15)

To contextualize our results, our data both agree and diverge from previous studies of head movement in patients with post-concussion central vestibular deficit and other studies investigating peripheral vestibular dysfunction. Sessoms and colleagues(7) assessed head kinematics during walking in tactical-athletes with post-concussion vestibular deficit and found asynchrony and large group variability of head position in relation to the pelvic COM. We similarly observed substantially large group variability, a finding likely resulting from heterogeneity of movement strategies employed during walking in the Vestibular group. Mijovic and colleagues(16) studied head movement in individuals with unilateral peripheral vestibular deficit during a 15-meter walking task using movement sensors and found no kinematic differences compared to healthy controls. This is likely a function of severity of impairment, with our sample having substantially higher DHI scores compared to those reported by Mijovic and colleagues.(16) While Pozzo and colleagues(17) found that individuals with bilateral peripheral vestibular deficit demonstrated no significant differences in head motion during walking measured with video analysis, they similarly observed greater variability in movement. Disparity between our findings and those previously reported are likely a function of lesion type and severity of symptoms.

### 4.2. Body mechanics

Basford and colleagues(18) studied body COM kinematics in individuals with traumatic brain injury with similar DHI scores and found that body COM excursion and velocity were significantly diminished in the AP and increased in the ML. These findings were attributed to decreased walking velocity and step length,(18) spatiotemporal alterations previously observed in this clinical population.(18,19) The supposition of pelvic excursion as a function of spatiotemporal parameters is supported by our findings. While we similarly saw increased mediolateral and vertical pelvic motion and velocity, we did not observe any differences in anteroposterior parameters. Since gait speed was standardized, the increased mediolateral pelvic excursion was likely attributed to the wider step width, a finding likely attributed to the neurological deficit.

The pelvic excursions and velocities observed in our study agree with a few previously-reported findings. Catena and colleagues(20) studied pelvic COM kinematics during over-ground walking in individuals post-concussion and found that pelvic COM velocity were significantly increased in the ML, with significantly lower COM velocity in the AP. They did not find any significant differences in pelvic excursion. (20) In a later study, Catena and colleagues(21) found no significant differences in COM kinematics between post-concussion patients and healthy controls during single-task walking up to 28 days post injury. Parker and colleagues(22) found that while AP displacement and velocity were not significantly different between post-concussion patients and healthy controls up to 28 days following concussion, there was a significantly diminished maximum separation difference between AP COM and center of pressure indicating a more conservative strategy for stability. Reciprocal arm swing has been postulated to be a mass dampener that mitigates trunk torsion and head yaw during walking.(23) While we did not measure upper extremity kinematics, it is plausible that asynchronous arm swing likely contributed to alteration in pelvic and head mechanics in this study.

### 4.3 Clinical and research implications

Clinical and instrumented measures of head motion during gait may garner additional information regarding central neurological mechanisms and may be an important clinical correlate in the assessment of the balance system during function. Based on heterogeneity of clinical presentation following concussion, diminished head motion may be a useful measure when diagnosing, prognosticating, and guiding treatment in this population. Physical examination should include assessment of cervical motion, cervicogenic dizziness, dynamic visual acuity, gaze stabilization, subjective visual vertical, static balance using the SOT, and gait. Identified deficits to include diminished head kinematics during gait should be addressed during rehabilitation to allow the patient to reach their full potential. Further research is needed to investigate the relationship between head motion and optical tracking and stabilization and how these change in response to vestibular rehabilitation.

### 4.4. Limitations

There are limitations to this study. This was a cross-sectional study, so cause and effect relations cannot be determined. The study sample consisted of young adult tactical athletes with a high level of physical function prior to injury, hence generalizability is likely limited to individuals of similar age and physical function. Kinematic measurements were focused on assessment of the pelvic and head COM during walking. Three-dimensional kinematic evaluation of head, arm, trunk, and lower extremity kinematics were not possible based on the marker set used in this study. Future study should consider the intersegmental coupling relationships that can further elucidate these findings.

## 5. Conclusion

Patients with post-concussion vestibular deficit demonstrated a more constrained head and pelvic movement strategy during gait compared with the Control group, a finding that is likely attributed to a neurological impairment of visual-vestibular-somatosensory integration. Clinicians and researchers should consider assessment of head and neck movement during gait following concussion, as this information may provide greater insight to impaired peripheral and central vestibular-visual-somatosensory mechanisms.

## Data Availability

Due to the nature of this research, participants of this study did not agree for their data to be shared publicly, so supporting data is not available.

## Role of the funding source

None.

## Declaration of Competing Interest

Authors Jacob VanDehy, Dawn M. Bodell, and Kim R. Gottshall was employed by the company Leidos, Inc. The remaining authors declare that the research was conducted in the absence of any commercial or financial relationships that could be construed as a potential conflict of interests.

## Author contributions

John J. Fraser contributed to study design, data analysis and interpretation, and drafting this manuscript. Jacob VanDehy contributed to data collection and analysis. Dawn M. Bodell contributed to study design, participant recruitment, data collection, and interpretation of findings. Kim R. Gottshall contributed to study design, data interpretation, and drafting this manuscript. Pinata H. Sessoms contributed to study design, procurement of funding, data collection, and data interpretation. All authors have reviewed and approved this manuscript.

## Notes

Funding: This work was supported by the U.S. Navy Bureau of Medicine and Surgery under work unit no. N1703.

### Competing Interest Statement

The authors have declared no competing interest.

### Funding Statement

This work was supported by the U.S. Navy Bureau of Medicine and Surgery under work unit no. N1703.

### Author Declarations

The study protocol was approved by the Naval Health Research Center Institutional Review Board in compliance with all applicable Federal regulations governing the protection of human subjects. Research data were derived from an approved Naval Health Research Center Institutional Review Board protocol, number NHRC.2015.0010. All participants provided informed consent.

### Summary of Updates

Corrected and reconciled the number of participants in each group in the abstract, flow diagram, and in the text of the methodology section. Updated disclosure for Leidos affiliated authors.

